# Plasma cfDNA analysis can be facilitated by membrane based filtration of whole blood regardless of additive

**DOI:** 10.1101/2023.07.05.23292260

**Authors:** Amanda N. Marra, Dirk van den Boom, Mathias Ehrich, Christopher K. Ellison

## Abstract

Cell-free DNA (cfDNA) has emerged as a pivotal biomarker for various noninvasive screening applications such as noninvasive prenatal screening, minimum residual disease monitoring, and transplant rejection prediction. However, the efficacy of these applications is heavily reliant on the detection of minority nucleic acid species amidst a vast background of host DNA. Therefore, it is crucial to prevent the dilution of the desired cfDNA signal with genomic DNA originating from white blood cell (WBC) degradation. The most commonly adopted sample collection methods prevent WBC lysis by either adding a stabilizing agent to the collection tube, like Streck, or through immediate centrifugation following the collection of whole blood. In this study, we propose an alternative for plasma sample collection that involves membrane based filtration of plasma from cellular blood components at the point of collection. Our findings demonstrate that cfDNA recovered using this method performs equally or better compared to conventional methods and enables the implementation of novel and more accessible blood collection procedures.

## 1 Introduction

Plasma cell-free DNA (cfDNA) is highly fragmented circulating DNA that is most often released from apoptotic cells of the hematopoietic system (1, 2). cfDNA is abundant in peripheral blood, where it can easily be retrieved from the plasma of a blood sample. In addition to hematopoietic cells, cfDNA can originate from many different tissue sources in the body (1, 3-5), which has made cfDNA a useful biomarker for many clinical screening assays. Specific clinical applications include detection of cf fetal DNA released from the placenta in non-invasive prenatal screening (NIPS) (6-13), from solid tumors in minimal residual disease detection (14-18), and from donor organs in transplant rejection monitoring (19, 20).

The majority of these noninvasive clinical applications are designed to detect a minority species of cfDNA amongst a vast background of circulating host DNA. In a whole blood sample, there is about 1000 fold more genomic DNA (gDNA) in white blood cells (WBCs) than the amount of cfDNA in plasma. Therefore, it is crucial to prevent the release of gDNA from WBCs to avoid dilution of the minority cfDNA species. Traditionally, blood samples for noninvasive applications are collected from patients intravenously by a licensed phlebotomist and plasma is isolated via centrifugation. Centrifugation physically separates whole blood into cellular and plasma components, and because WBCs are subject to degradation, centrifugation of the sample must be performed nearly immediately after collection (21-27). In order to extend the processing time, a preservative such as Streck must be added to the whole blood sample to prevent WBC lysis and stabilize cfDNA for downstream applications (24, 25, 28, 29).

While the use of Streck blood collection tubes has been implemented in clinical settings, the reliance on phlebotomy limits the application. Barriers such as finding an in-network provider or travel to and from the clinic for a blood draw still pose a burden on patients (30). Furthermore, the amount of cfDNA in plasma is relatively dynamic and can be influenced by both intrinsic and extrinsic factors (31). Therefore, it is unlikely that a single snapshot of cfDNA has the same predictive power as a series of collections taken over a period of time. For example, monitoring organ health in allograft recipients or remission in cancer patients may benefit from frequent sample collections for cfDNA testing to improve clinical accuracy. Because of the logistics involved in cfDNA collections, studies to further investigate these phenomena might simply be impractical. To increase the accessibility and reach of cfDNA research and its applications, a sample collection method is required that eliminates the need for phlebotomy, such as self sampling of capillary blood, and combines it with a method that prevents WBC contamination. While self sampling through capillary blood collection is ubiquitously available, adding a stabilizing agent is impractical since most are formaldehyde based and should not be used close to an open wound or within the home.

We have devised a sample collection device (SCD) that relies on membrane based filtration to isolate plasma from whole blood samples. With this concept, capillary blood is applied onto a filtration membrane which physically separates cellular blood components from the plasma. The filtered plasma is then collected in a sealed vial, securing the plasma for transportation. Here, we evaluate the performance characteristics of this alternative sample collection method and compare it directly to standard cfDNA collection methods. In particular, we examine if cfDNA recovered from the SCD is free from WBC contamination and stable over a week at room temperature. Additionally, we determine whether our proposed sample collection method is suitable for self sample collection by patients and if the resulting cfDNA shares the same characteristics as cfDNA recovered from standard methods of blood sample collection.

## 2 Materials and Methods

### 2.1 Ethics statement

All sample collection and use for research was approved by the WCG Independent Review Board. Venous samples from nonpregnant individuals were performed by a licensed phlebotomist under IRB protocol #Juno-2017-0001. Commercial samples were de-identified from all patient health information and analyzed under IRB protocol #202304RPS. All commercial samples were capillary self-collections performed at home with the SCD (Video S1).

### 2.2 Male sample collection

Venous blood from a single male donor was drawn sequentially into 6 mL BD Vacutainer® EDTA (BD #367863) and Streck Cell-free DNA (Omaha, NE, USA) blood collection tubes, then separated into cellular components and plasma via centrifugation at room temperature. The centrifuge protocol used for whole blood separation was as follows: (1) spin at 300 RCF for 20 minutes with the deceleration brake set to 0, (2) recover plasma in a sterile conical tube, (3) spin recovered plasma at 2,000 RCF for 10 minutes with the deceleration brake set to 4, and (4) recover plasma in a sterile conical tube for storage. The recovered plasma samples were kept at room temperature under 30 minutes while female blood was collected.

### 2.3 Nonpregnant female sample collection

Venous blood from a single female donor was drawn sequentially into 6 mL EDTA and Streck collection tubes. Immediately following collection, 5 mL of blood was transferred to a sterile 15 mL conical tube. To mimic a minority male fetal signal, 500 μL of the corresponding male plasma was added directly to the 5 mL of female blood so that EDTA male plasma was added to EDTA female blood and Streck male plasma was added to Streck female blood. The mixtures were inverted to mix, then transferred to microcentrifuge tubes and SCDs at 100 μL each. All SCDs were activated to separate plasma from blood. At days 0, 1, 4, and 8 after sample collection, plasma was recovered from the SCDs. Concurrent to plasma recovery from the SCDs, whole blood aliquots in microcentrifuge tubes were separated into plasma by centrifugation and the final plasma was collected (Figure 1).

**Figure 1:**
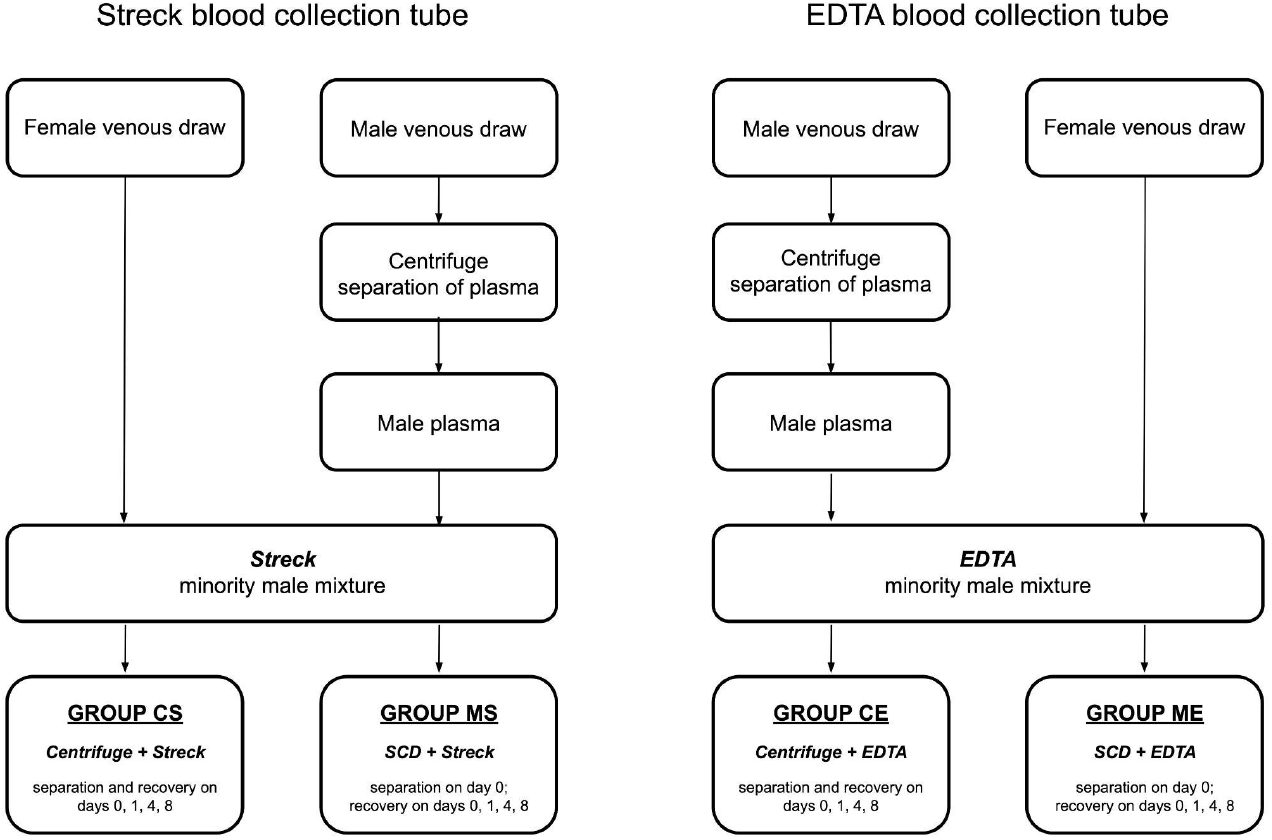
Schematic of experimental setup. Venous draws were collected from a single male individual and a single nonpregnant female individual. SCD = sample collection device.

### 2.4 cfDNA extraction

The MagMAX™ Cell-Free DNA Isolation Kit (Applied Biosystems™, ThermoFisher) was used to extract cfDNA from plasma according to the manufacturer’s instructions. Samples were processed on an automated liquid handler using custom protocols. Purified cfDNA was eluted in MagMAX™ Cell-Free DNA Elution Solution and stored at 4C.

### 2.5 Next-generation sequencing and downstream analysis

Library preparation was performed on an automated liquid handler with Aequitas Library Prep reagents and purification beads (Juno Diagnostics) using custom protocols. Library concentrations were quantified using the Qubit™ 1X dsDNA High Sensitivity Assay (Invitrogen, ThermoFisher). Then, libraries were normalized and pooled for sequencing on an Illumina NextSeq 2000. Sequencing results were analyzed through a Juno Diagnostics proprietary pipeline for bioinformatic analysis.

### 2.6 Real-time PCR

Real-time PCR primers and probes were designed to target *DXZ* for total genomes and *DYZ* for male genomes (32). cfDNA, primers and probe were added to Platinum™ Quantitative PCR SuperMix-UDG w/ROX (Invitrogen, ThermoFisher) to make 50 μL reactions. Real-time PCR was performed on QuantStudio 3 and 6 Real-Time PCR instruments. Data was analyzed with the QuantStudio software to determine DNA quantity and then converted to total or male genomes/μL plasma.

### 2.7 Data visualization and statistical analysis

Data was analyzed and visualized in R (version 4.2.2) on RStudio (version 2022.07.0) with the following packages: ggplot2 (version 3.3.6), tidyverse (version 1.3.2), splitstackshape (version 1.4.8), googlesheets4 (version 1.0.0), and patchwork (version 1.1.2). Statistical analysis was performed for each condition in relation to day 0 using a Wilcoxon test, where p-values of p < 0.05 are significant.

## 3 Results

We used a contrived blood sample that contained a minority fraction of male cfDNA to evaluate the total amount of DNA recovered as well as the characteristics of the recovered DNA. We investigated four different methods of blood collection: blood collection in Streck followed by separation of plasma via either centrifugation (Group CS) or SCD (Group MS) and blood collection in EDTA followed by separation of plasma via either centrifugation (Group CE) or SCD (Group ME) (Figure 1). This setup necessitates plasma separation to be performed on day 0, 1, 4, and 8 for groups CS and CE, while for groups MS and ME plasma separation occurred at the time of collection (detail in methods).

### 3.1 Quantification of total DNA amounts and gDNA contamination

In the reference groups CS and CE, the median amount of DNA recovered on day 0 was 4.3 and 4.2 genomes/μL plasma, respectively (Table S1). The median amount of total DNA recovered following membrane based filtration of plasma from whole blood was 2.5 genomes/μL plasma in group MS and 3.2 genomes/μL plasma in group ME (Table S1). This loss of DNA is in line with expectations for membrane based filtration and likely due to surface interactions of cfDNA with the hydrophilic polysulfone membrane.

To compare results across all timepoints and conditions, we normalized the total DNA amount to the median amount for each group on day 0. Neither of the membrane based groups (MS and ME) showed a statistically significant change in total DNA over the course of the study (Table S1, Figure 2A, S1). In the centrifugation based groups, we observed a statistically significant decrease for samples collected with the Streck additive (−14%) and a statistically significant increase for the samples collected in EDTA (+17500%). The increase in centrifuged EDTA collected blood is due to WBC degradation, and this has been well described in the literature (21-27). After 24 hours, the CE group already showed a 2.8 fold increase in total DNA, which ultimately rose to more than 170 fold increase by day 8 (Table S1, Figure 2A). The decrease observed in centrifuged Streck collected blood (Table S1, Figure S1) is likely due to increased crosslinking of cfDNA with proteins, making it less accessible to the DNA extraction method used in this study (28, 33).

**Figure 2:**
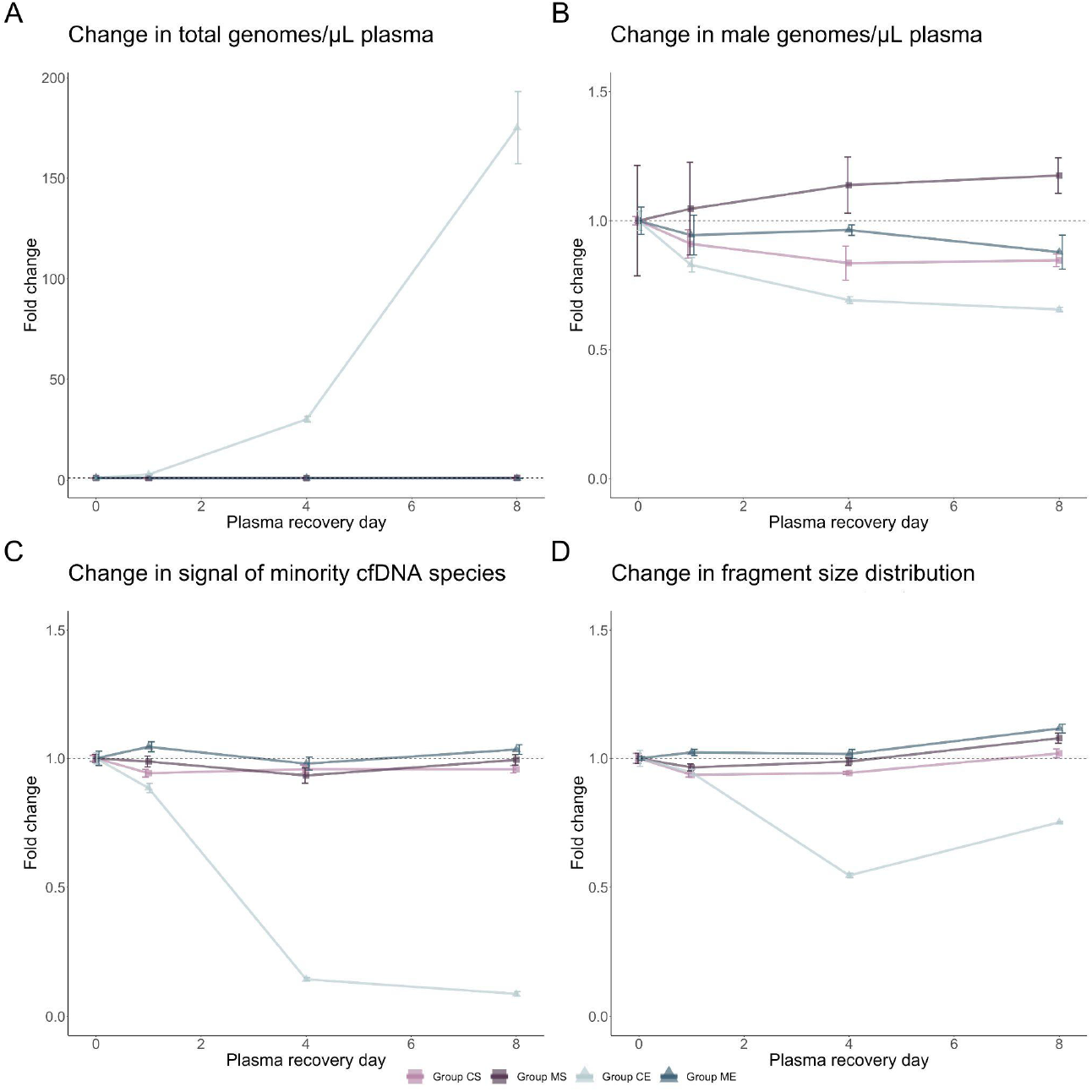
Membrane based filtration of plasma reduces gDNA contamination and preserves cfDNA signal. (A) Change in total genomes/μL plasma. (B) Change in male genomes/μL plasma. (C) Change in signal of minority cfDNA species. (D) Change in fragment size distribution. For A-D, median values are normalized to the median at day 0 for each condition. Group CS (centrifuge + Streck) is represented by light purple and square points, group MS (SCD + Streck) is represented by dark purple and square points, group CE (centrifuge + EDTA) is represented by light blue and triangle points, and group ME (SCD + EDTA) is represented by dark blue and triangle points. Standard error is shown for each condition at each recovery day.

### 3.2 Quantification of minority cfDNA species

The amount of male genomes/μL plasma remained stable for groups that used membrane based filtration (MS and ME) (Table S1, Figure 2B). Similar to the observed decrease in total DNA amount for Streck standard group CS, this group displayed a 20% decrease in the median amount of male genomes/μL plasma over the span of 8 days (Table S1). As discussed above, this decrease is most likely a consequence of crosslinked cfDNA and proteins impacting DNA extraction efficiency. However, in group CE, there was a statistically significant decrease in the amount of male genomes/μL plasma equating to about 40% loss by day 8 (Table S1, Figure 2B). This loss may be a consequence of low-level residual DNase activity in EDTA plasma samples (34). These data suggest that membrane based filtration of plasma from whole blood stabilizes minority cfDNA species within the plasma for at least eight days storage time.

### 3.3 cfDNA characteristics are preserved with membrane based filtration

Besides simple assessment of cfDNA quantity, a variety of other cfDNA characteristics have been used in clinical and research applications. cfDNA stability and amount may have a significant and measurable impact on potential downstream applications such as detecting minority signal abundance (6-20) and cfDNA fragmentomics (4, 35-38). Both signal abundance and fragment distribution are commonly assayed using Next Generation Sequencing (NGS) approaches.

To evaluate the characteristics of cfDNA recovered from membrane based filtration with the SCD, we used NGS to measure the representation of the minority species of cfDNA in our contrived cfDNA samples. Sequencing data shows that cfDNA fragments from the male minority species are present at a stable rate from day 0 to day 8 for groups CS, MS and ME (TableS1, Figure 2C). The EDTA samples that were separated by centrifugation (group CE) showed a progressive decline in male cfDNA representation with a reduction of 91% by day 8 relative to day 0 (Table S1, Figure 2C). The representation of male DNA in general was concordant between sequencing based measurements and real-time PCR based measurements (Figure S2). In many implementations, the magnitude of this change in minority signal will fundamentally alter the results of the assay.

Fragmentomic approaches have been used as a diagnostic tool to parse the tissue source of cfDNA (4, 35-38). To compare the fragment size profiles of samples from standard methods of cfDNA recovery (groups CS and CE) to membrane based filtration (groups MS and ME), we calculated the ratio of small (100 - 150 bp) to large (151 - 220 bp) DNA fragments using a previously reported method (36). The ratio of small to large fragments does not change much over the span of 8 days in groups CS (+0%) and MS (+9%). For group ME we observed a slight increase in smaller DNA, possibly due to residual DNase activity (+14%) (Table S1, Figure 2D). In group CE, the proportion of large fragments significantly increased over the course of the study (−29% in ratio of small to large fragments) (Table S1, Figure 2D), consistent with previous indication of WBC lysis.

### 3.4 Patient applicability

To assess the feasibility of membrane based filtration in practical applications, we analyzed self-collected capillary blood samples that were sent in for a commercial fetal sex test. This set included over 3100 samples collected by pregnant women using the SCD in a home setting. The separated plasma was shipped to the lab using commercial carriers and accessioned upon receipt. We observed that 99% of the devices yielded sufficient material to be assayed for cfDNA whole genome sequencing. These data suggest that membrane based filtration of plasma from whole blood preserves the cfDNA signal and provides a robust pre-analytical solution for cfDNA assays.

## 4 Discussion

The explosion in scientific literature focusing on cfDNA since the year 2000 (27) highlights the importance of using cfDNA as a biomarker. As the popularity of noninvasive screening continues to grow, optimization of the pre-analytical sample collection process is a crucial step towards downstream assay success. Here, we show that the quality of plasma cfDNA recovered from whole blood samples following membrane based filtration is equivalent to standard collection methods. Most importantly, even in the absence of stabilizing agents, the physical separation of WBCs from plasma at the time of collection prevents gDNA contamination and dilution of cfDNA signal (Figure 2A, 2C; Table 1, S1). Furthermore, membrane based filtration does not impact the detection of minority constituents of cfDNA (Figure 2B, 2C; Table 1, S1). These observations have been shown to be stable for at least 8 days (up to 14 days, internal data not shown). These results support the use of membrane based filtration of plasma from whole blood in clinical and research applications, eliminating the need for phlebotomy and increasing the flexibility of sample collection and sample logistics.

**Table 1:** Summary of sample characteristics observed from day 0 to day 8.

| Condition | WBC lysis | Dilution of minority cfDNA species | Loss of minority cfDNA signal |
| --- | --- | --- | --- |
| centrifuge + EDTA | + | + | + |
| centrifuge + Streck | - | - | - |
| SCD + EDTA | - | - | - |

In current clinical practice, obtaining whole blood samples often requires an intravenous draw performed by a licensed phlebotomist. While phlebotomy infrastructure is reasonably well developed in urban areas, access is more restricted in rural regions. Also, a recent report shows that the time required for an appointment is around 120 minutes (30), which places a significant burden on patients, especially when frequent draws are necessary. Furthermore, more than 5% of the population present challenges to successful phlebotomy collection and could benefit from alternative collection modalities (39). A device like the SCD unlocks a multitude of possibilities in clinical applications and research. For example, it can enable people in rural areas to provide a capillary self collected sample without major disruptions to their workday for routine testing such as NIPS. Additionally, because the SCD does not require stabilizers to prevent WBC lysis, the risk of exposure to potentially toxic chemicals is eliminated. Without the need for a phlebotomist and preservatives to collect blood samples, membrane based filtration of plasma from whole blood enables far more interesting scenarios of frequent testing in monitoring applications. For example, in allograft rejection monitoring, typically only a few timepoints are measured. It is currently unknown if weekly monitoring would enhance the predictive power of such an assay by not only having a snapshot of information available but also the dynamic changes over time. This is a huge unanswered question which can now be easily addressed.

The easy-to-use, portable, and stable method of plasma collection through membrane based filtration of whole blood offers numerous opportunities for scientific research in various areas. One area that can benefit are investigations of the impact of exercise on cfDNA levels. Typically, these studies are conducted in highly controlled laboratory settings, such as at an university exercise laboratory. With the increased flexibility provided by membrane based filtration, it is possible to easily perform sample collection at the point of interest. For example, study participants can sample over greater time periods while running an amateur race or engaging in physically demanding activities such as hikes or mountaineering. While it is widely accepted that exercise does increase the amount of circulating cfDNA, the effect of other intrinsic (i.e circadian rhythm) and extrinsic (i.e. stress, diet, and alcohol intake) factors remains more nebulous (31). Increasing sample frequency will greatly contribute to a better understanding of how these factors affect cfDNA dynamics. For instance, all day sampling under various conditions can uncover cfDNA patterns that would impact the outcomes of noninvasive assays. This would be particularly valuable in assays designed to detect minority species of cfDNA. In NIPS, for example, the detection of fetal cfDNA can be positively or negatively impacted by the amount of maternal circulating cfDNA at the time of sample collection.

By eliminating the need for phlebotomy and preservatives, membrane based filtration allows for whole blood sample collection at any time and location. This method not only enhances clinical accessibility by providing both scheduling and location flexibility for sample collection, but it also enables basic research projects to uncover deeper insights into cfDNA dynamics. In summary, the adoption of membrane based filtration of plasma from whole blood combined with capillary self-sampling has the power to revolutionize both basic cfDNA research and clinical applications.

## Data Availability

All data produced in the present study are available upon reasonable request to the authors.

## 5 Figure Legends

**Figure S1:**
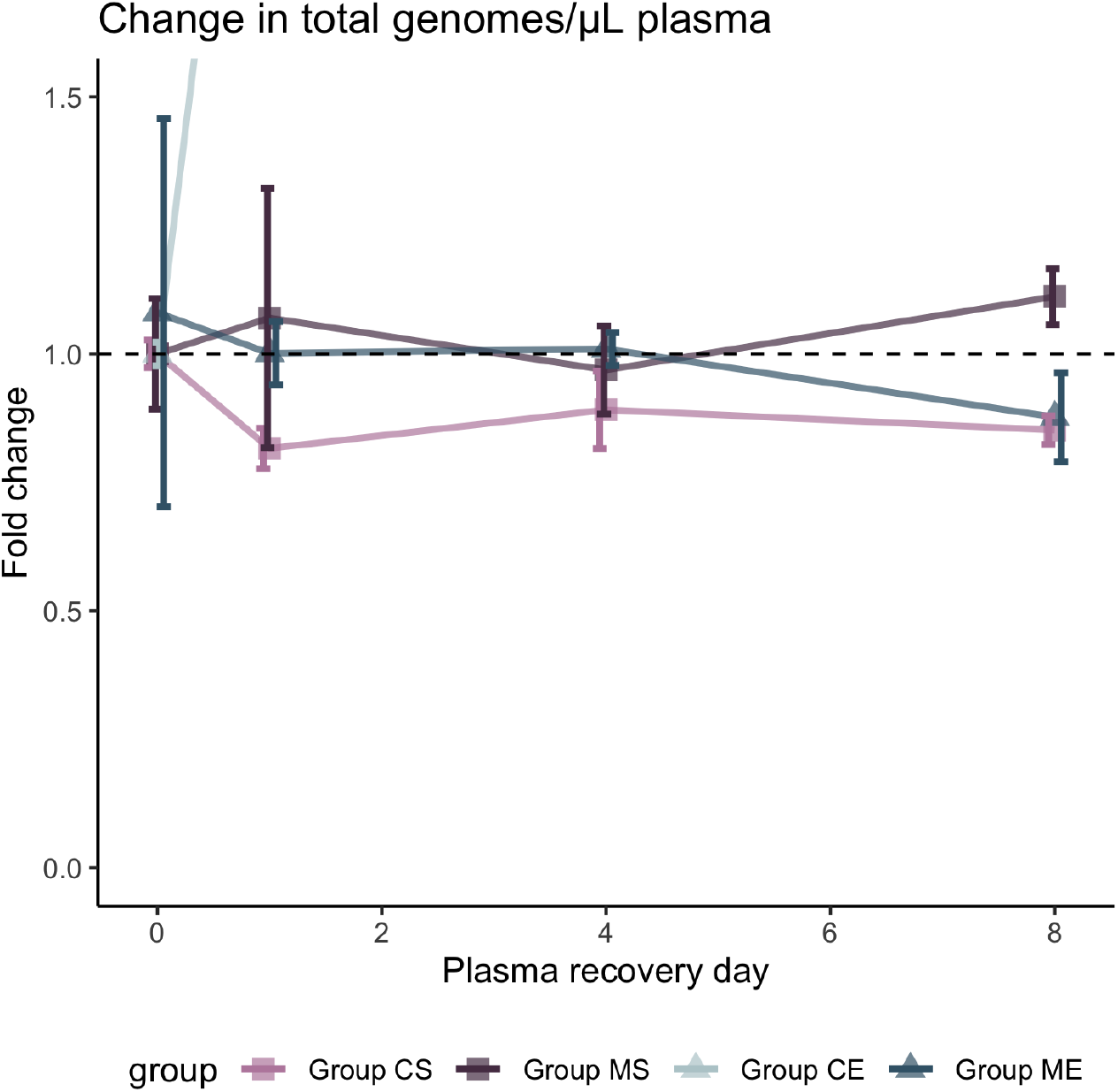
Change in total genomes/uL plasma over 8 days. For each condition, median values are normalized to the median at day 0. Group CS (centrifuge + Streck) is represented by light purple and square points, group MS (SCD + Streck) is represented by dark purple and square points, group CE (centrifuge + EDTA) is represented by light blue and triangle points, and group ME (SCD + EDTA) is represented by dark blue and triangle points. Standard error is shown for each condition at each recovery day. The y-axis is scaled to show distribution of group CS, MS, and ME.

**Figure S2:**
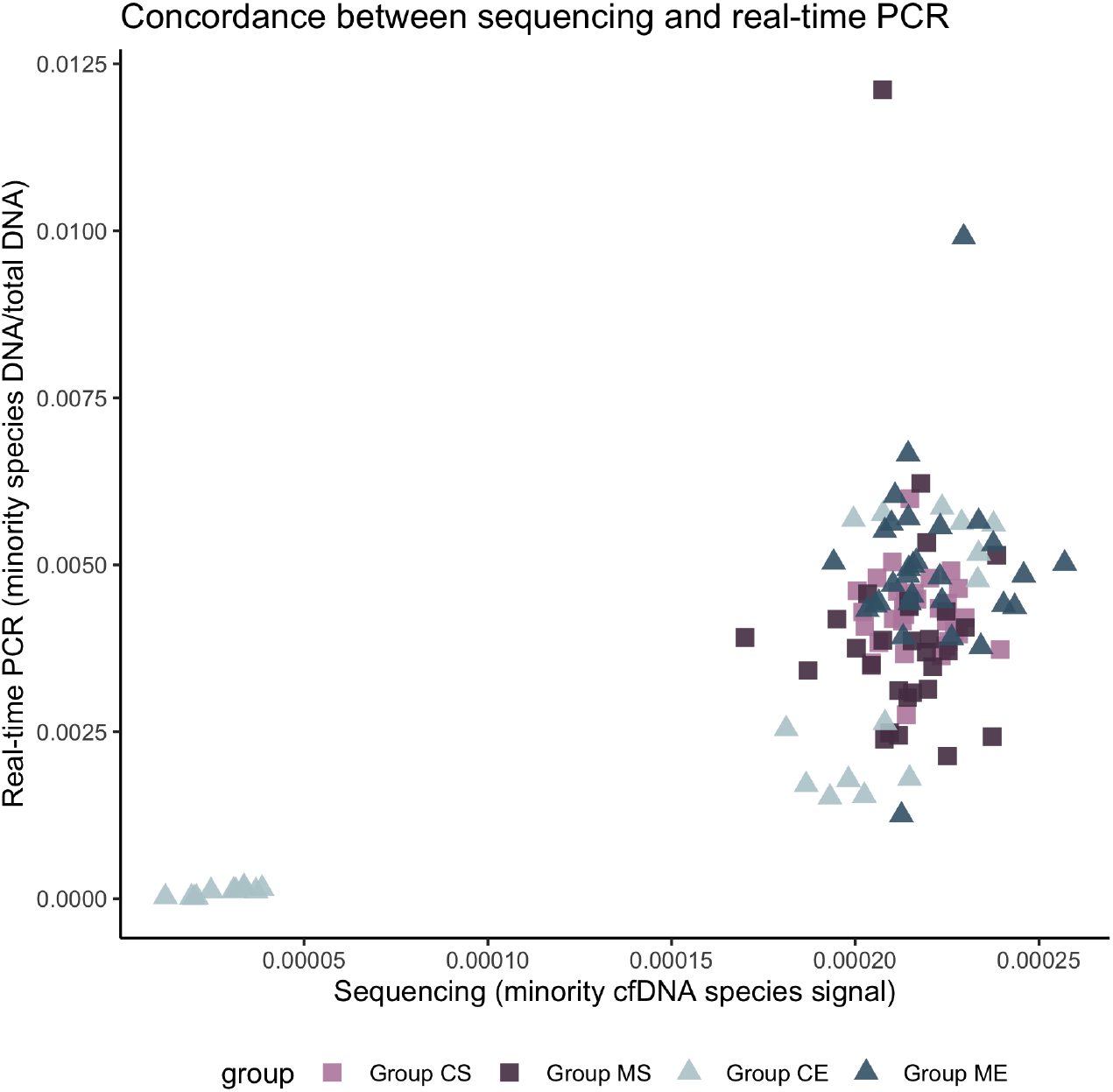
Concordance between sequencing measurements and real-time PCR measurements. Group CS (centrifuge + Streck) is represented by light purple square points, group MS (SCD + Streck) is represented by dark purple square points, group CE (centrifuge + EDTA) is represented by light blue triangle points, and group ME (SCD + EDTA) is represented by dark blue triangle points.

5.5 Video S1: Capillary self-collection with the Sample Collection Device (SCD). After washing hands and thoroughly wiping the non-dominant ring or index finger with ethanol, a lancet is used to puncture the finger. The first drop of blood is wiped away, then blood is collected in an EDTA-coated minivette. Once full, the minivette is inverted and the blood is dispensed onto the sample collection pad. Following the application of pressure, plasma passes through the filter under the sample collection pad and is collected in a plastic vial. The shipping sleeve then moves the vial into the final position underneath the foil seal, securing the plasma for shipment.

## 6 Tables

**Table S1:** Data summary

| characteristic | group | day 0 median | day 1 median | day 4 median | day 8 median | day 0 vs 1 (p-value) | day 0 vs 4 (p-value) | day 0 vs 8 (p-value) |
| --- | --- | --- | --- | --- | --- | --- | --- | --- |
| total genomes | CS | 4.3 | 3.5 | 3.8 | 3.7 | 0.0104 | 0.3823 | 0.0047 |
| total genomes | MS | 2.5 | 2.7 | ** 2.5 | 2.8 | 1.0000 | 1.0000 | 0.6454 |
| total genomes | CE | * 4.2 | * 11.6 | 127.1 | 737.3 | 0.0006 | 0.0003 | 0.0003 |
| total genomes | ME | 3.2 | 3.0 | 3.0 | * 2.6 | 0.6454 | 0.5737 | 0.4634 |
| male genomes | CS | 0.10 | 0.09 | 0.08 | 0.08 | 0.2345 | 0.0148 | 0.0011 |
| male genomes | MS | * 0.05 | 0.05 | ** 0.06 | 0.06 | 0.3357 | 0.9452 | 0.7789 |
| male genomes | CE | * 0.13 | * 0.11 | * 0.09 | * 0.08 | 0.0111 | 0.0006 | 0.0006 |
| male genomes | ME | * 0.09 | 0.08 | 0.08 | * 0.08 | 0.3357 | 0.1893 | 0.1282 |
| minority male fraction | CS | 0.16 | 0.15 | 0.15 | 0.15 | 0.0954 | 0.5557 | 0.2888 |
| minority male fraction | MS | 0.15 | 0.15 | 0.14 | 0.15 | 0.4183 | 0.0435 | 0.5680 |
| minority male fraction | CE | 0.16 | 0.14 | 0.02 | ** 0.00 | 0.0096 | 0.0007 | 0.0074 |
| minority male fraction | ME | 0.15 | 0.16 | 0.15 | 0.16 | 0.3638 | 0.1640 | 0.5999 |
| small to large fragment ratio | CS | 0.24 | 0.22 | 0.22 | 0.24 | 0.0353 | 0.0457 | 0.0738 |
| small to large fragment ratio | MS | 0.23 | 0.22 | 0.23 | 0.25 | 0.1403 | 1.0000 | 0.0207 |
| small to large fragment ratio | CE | 0.21 | 0.20 | 0.11 | 0.15 | 0.0180 | 0.0009 | 0.0009 |
| small to large fragment ratio | ME | 0.21 | 0.22 | 0.22 | 0.24 | 0.0513 | 0.2254 | 0.0009 |

For each condition, n = 8 unless annotated (\* n = 7; \*\* n < 7)

## 8 Author Contributions

A.N.M., C.K.E, D.vdB. and M.E. designed research; A.N.M. performed research; A.N.M., C.K.E, D.vdB. and M.E. analyzed data; and A.N.M., C.K.E, and M.E.wrote the paper.

## 10 Acknowledgments

We would like to thank Nicole Razon, AAMA, CPT I, for performing venous blood collections.

